# On Cross-ancestry Cancer Polygenic Risk Scores

**DOI:** 10.1101/2021.02.24.21252351

**Authors:** Lars G. Fritsche, Ying Ma, Daiwei Zhang, Maxwell Salvatore, Seunggeun Lee, Xiang Zhou, Bhramar Mukherjee

## Abstract

Polygenic risk scores (PRS) can provide useful information for personalized risk stratification and disease risk assessment, especially when combined with non-genetic risk factors. However, their construction depends on the availability of summary statistics from genome-wide association studies (GWAS) independent from the target sample. For best compatibility, it was reported that GWAS and the target sample should match in terms of ancestries. Yet, GWAS, especially in the field of cancer, often lack diversity and are predominated by European ancestry. This bias is a limiting factor in PRS research. By using electronic health records and genetic data from the UK Biobank, we contrast the utility of breast and prostate cancer PRS derived from external European-ancestry-based GWAS across African, East Asian, European, and South Asian ancestry groups. We highlight differences in the PRS distributions of these groups that are amplified when PRS methods condense hundreds of thousands of variants into a single score. While European-GWAS-derived PRS were not directly transferrable across ancestries on an absolute scale, we establish their predictive potential when considering them separately within each group. For example, the top 10% of the breast cancer PRS distributions within each ancestry group each revealed significant enrichments of breast cancer cases compared to the bottom 90% (odds ratio of 2.81 [95%CI: 2.69,2.93] in European, 2.88 [1.85, 4.48] in African, 2.60 [1.25, 5.40] in East Asian, and 2.33 [1.55, 3.51] in South Asian individuals). Our findings highlight a compromise solution for PRS research to compensate for the lack of diversity in well-powered European GWAS efforts while recruitment of diverse participants in the field catches up.

## Introduction

Translating findings from genome-wide association studies (GWAS) to clinical utility in terms of complex trait prediction is a major milestone in genetics research [1]. This is especially important for traits whose estimated heritability was reported to be high. However, the identified common single nucleotide polymorphisms (SNPs) seldom have deterministic consequences. While each identified common risk SNP contributes to the overall disease risk, by itself it is unlikely to predict a large degree of variation in a disease outcome and thus usually represents a poor predictor by itself. The combination of all risk SNPs into a polygenic risk score (PRS) is a popular approach to improve predictive power and can be valuable for risk stratification, i.e., the identification of a small subset of a population with extreme PRS values that is at higher risk to develop a disease [1].

The discovery of risk SNPs through GWAS often depends on very large sample sizes of genotyped data (hundreds of thousands of tag SNPs or more) especially if one aims to capture a large fraction of the SNP heritability [2-4]. Until recently, GWAS of this scale were either exclusively or predominantly based on European populations, trailed by Asian populations, while all other ancestry groups comprised less than 5% [5]. The resulting bias in published GWAS results [6] is passed on to the development and application of PRS for many complex traits and despite current efforts to increase diversity in genetics research will likely continue in the foreseeable future [6].

The lack of portability of PRS across populations with different ancestry compositions is known and usually attributed to differences in causal variants, linkage disequilibrium (LD) patterns, allele frequencies, and effect sizes [7, 8]. In addition, genotyping or imputation methods that were originally developed for European ancestry (EA) studies can amplify such differences [7, 8].

There are several examples of studies that explore PRS constructed using GWAS results from different ancestry groups. Belsky *et al*. [9] constructed an obesity PRS based on EA-GWAS and found that it performed poorly individuals of African American compared to those of EA.[9] Grinde *et al*. [10] assessed the performance of PRS based on EA GWAS in a Hispanic/Latino population for three groups of traits: anthropometric measures, blood pressure, and blood count. The EA-based PRS performed well for anthropometric and blood count traits but performed poorly for blood pressure traits [10]. EA-based PRS for these quantitative traits also showed on average a 3.3-fold decrease in predictive performance in East Asian population when compared to the European population [11]. Others have demonstrated an association between PRS and genetic ancestry [12, 13]. Simply put, the literature cautions against the transferability of EA-based GWAS to other populations [5, 8]. Recently we have provided a catalog of more than 500 PRS for various cancer using EA-based GWAS [14]. However, there are little or no reports on the transferability of cancer PRS or whether these PRS can be used for other ancestries.

The UK Biobank Study (UKB) offers detailed questionnaire, electronic health record (EHR) and genetic data representing an excellent resource to study the influence of genetic risk factors on common complex disease. While predominantly European ancestry, it also includes over 20,000 participants of self-reported non-EA ancestry (reported as “ethnic groups”) [15] that can, together with genetically inferred ancestry information, be stratified into the four main ancestry groups: African, East Asian, European or South Asian ancestry (S1 Table). Thus, UKB offers the opportunity to evaluate the performance of PRS across various ancestry groups and to assess the transferability of EA-based cancer PRS.

To increase power for such an evaluation, we focus on two common cancer traits, breast and prostate cancer. Both of these traits offer several advantages for PRS explorations: high disease prevalence, large fraction of heritability already explained through known risk variants, low chance of phenotype misclassification, and available full summary statistics from very large, EA-based GWAS [16, 17].

## Results

We constructed cancer PRS specifically for the European subgroup of UKB individuals using two different approaches for each cancer trait: “GWAS hits PRS” is an effect-size weighted PRS based on a sparse set of GWAS hits (independent risk SNPs with P-value below 5×10^−8^) and “PRS-CS”, a Bayesian-regression-based PRS method that uses continuous shrinkage (CS) priors [18]. Relatively sparse sets of 334 and 377 SNPs were incorporated in the GWAS hits PRS for breast cancer and prostate cancer, respectively. By contrast the PRS-CS constructs integrated over 1.1 million SNPs for each of the two cancers.

What can be clearly seen in Fig 1 are the different distributions of PRS across the European, South Asian, African and East Asian ancestry groups that were statistically significantly different in group means by one-way ANOVA (P < 2.31×10^−141^; S2 Table). Both breast cancer PRS were on average higher in non-EA groups, whereas prostate cancer PRS were higher in African and lower in East and South Asian ancestry groups (Fig 1; S2 Table). These differences were pronounced for the PRS-CS-based PRS. This is likely a result of the summation of hundreds of thousands allele-frequency differences between ancestry groups compared to a few hundred for the GWAS hits PRS. Overall, this suggests that these PRS are not directly transferable, e.g., a high breast cancer PRS in EA individuals might fall into the lower PRS distribution of Africans ancestry individuals. This can also be observed when using a single PRS scale on the overall, heterogenous UK population, e.g., almost all African ancestry females have breast cancer PRS-CS scores above the population top 10% threshold while no East Asian ancestry male had a prostate cancer PRS-CS score above the population top 10% threshold (Fig 1).

**Fig 1.**
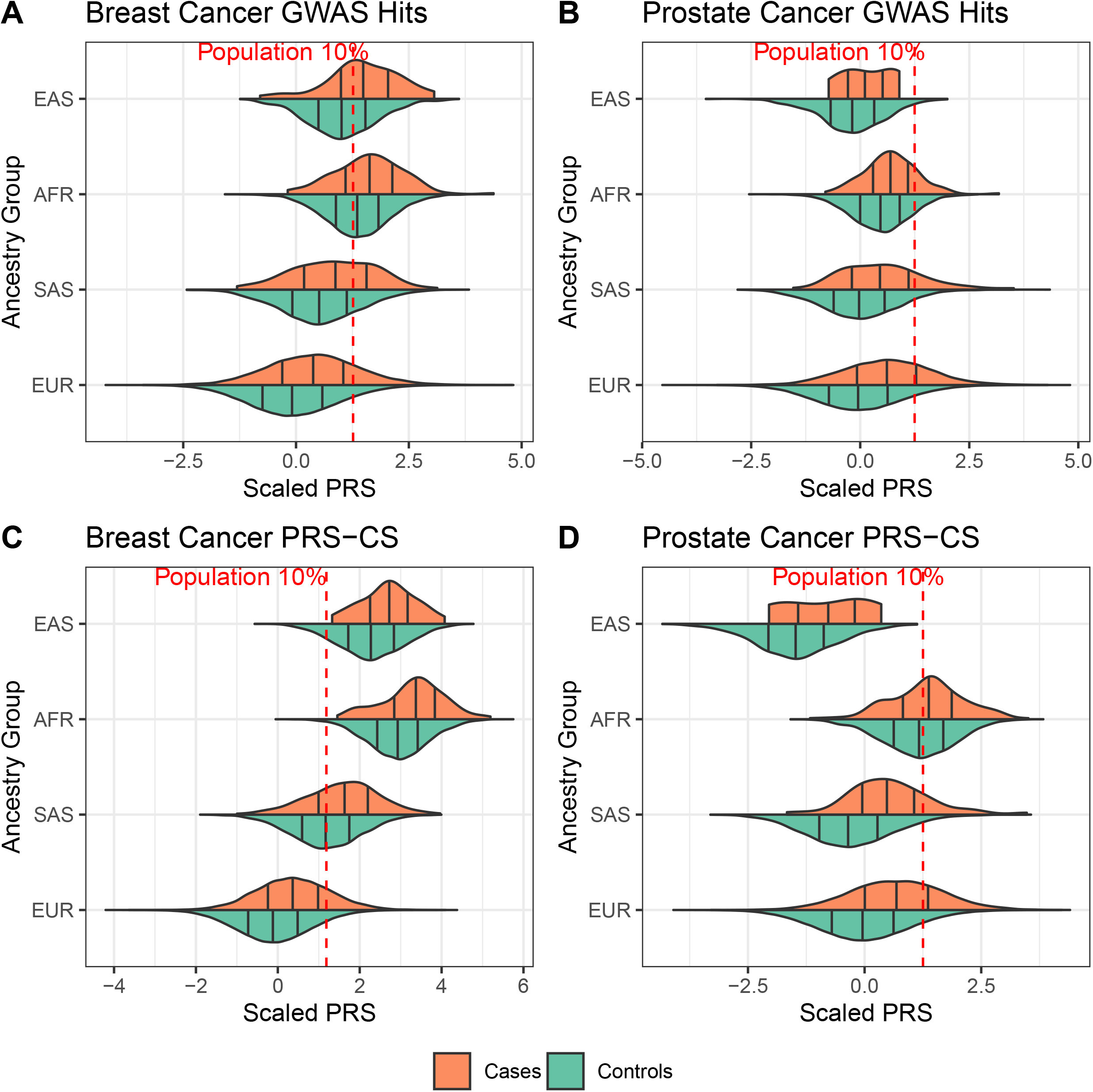
Violin plots of the breast and prostate cancer PRS distributions. Breast cancer (left) and prostate cancer (right) GWAS hit-based PRS (top) and PRS-CS-based PRS (bottom) stratified by ancestry group are shown. Black vertical lines indicate 25, 50, and 75% quantiles within the ancestry-specific case (orange) and control (green) distributions. Red lines indicate 10% quantiles of the corresponding UKB PRS distribution in all controls. Sample sizes for each sub-set can be found in Table 1.

Still, what is striking is the consistent right shift of the PRS distributions in cases compared to controls with each ancestry group (Fig 1). With exception of the small sample of East Asian prostate cancer cases (n = 7), all PRS were *significantly* associated with increased continuous ORs for their corresponding cancers when standardized to one standard deviation (S.D.) within each ancestry group (OR [per unit S.D.] ≥ 1.44, Table 1). Furthermore, all PRS also indicated satisfactory discriminative performance within each ancestry group (covariate-adjusted AUC [AAUC] > 0.589).

**Table 1.** Association and evaluation of cancer PRS across ancestry groups.

| GWAS Trait / Outcome | PRS Method (SNPs) | Ancestry Group | n Cases | n Controls | PRS Association |  | PRS Evaluation<br>AAUC<br>(95% CI) |
| --- | --- | --- | --- | --- | --- | --- | --- |
|  |  |  |  |  | Odds Ratio*<br>(95% CI) | P |  |
| Overall Breast Cancer | GWAS Hits (334) | EUR | 14109 | 214163 | 1.594<br>(1.567, 1.622) | 1.7x10<sup>-613</sup> | 0.628<br>(0.623, 0.632) |
|  |  | SAS | 149 | 3598 | 1.451<br>(1.231, 1.71) | 8.80x10<sup>-6</sup> | 0.603<br>(0.557, 0.651) |
|  |  | AFR | 116 | 3666 | 1.442<br>(1.199, 1.735) | 0.00011 | 0.6<br>(0.546, 0.65) |
|  |  | EAS | 45 | 1069 | 1.852<br>(1.385, 2.475) | 3.20x10<sup>-5</sup> | 0.676<br>(0.59, 0.757) |
|  | PRS-CS (1,120,410) | EUR | 14109 | 214163 | 1.771<br>(1.739, 1.803) | 5.4x10<sup>-857</sup> | 0.653<br>(0.648, 0.657) |
|  |  | SAS | 149 | 3598 | 1.656<br>(1.4, 1.958) | 4.00x10<sup>-9</sup> | 0.641<br>(0.594, 0.687) |
|  |  | AFR | 116 | 3666 | 1.761<br>(1.453, 2.134) | 7.90x10<sup>-9</sup> | 0.651<br>(0.598, 0.701) |
|  |  | EAS | 45 | 1069 | 1.761<br>(1.297, 2.39) | 0.00029 | 0.66<br>(0.585, 0.735) |
| Prostate Cancer | GWAS Hits (377) | EUR | 6561 | 182590 | 1.943<br>(1.894, 1.993) | 3.1x10<sup>-566</sup> | 0.68<br>(0.674, 0.687) |
|  |  | SAS | 51 | 4305 | 1.785<br>(1.389, 2.295) | 6.00x10<sup>-6</sup> | 0.652<br>(0.576, 0.726) |
|  |  | AFR | 144 | 2681 | 1.501<br>(1.254, 1.796) | 9.70x10<sup>-6</sup> | 0.615<br>(0.567, 0.66) |
|  |  | EAS | 7 | 622 | 1.63<br>(0.83, 3.205) | 0.16 | 0.619<br>(0.442, 0.811) |
|  | PRS-CS (1,120,596) | EUR | 6561 | 182590 | 2.14<br>(2.085, 2.197) | 4.2x10<sup>-711</sup> | 0.702<br>(0.695, 0.708) |
|  |  | SAS | 51 | 4305 | 2.383<br>(1.826, 3.111) | 1.70x10<sup>-10</sup> | 0.745<br>(0.684, 0.8) |
|  |  | AFR | 144 | 2681 | 1.325<br>(1.107, 1.586) | 0.0021 | 0.579<br>(0.527, 0.63) |
|  |  | EAS | 7 | 622 | 1.943<br>(1.005, 3.755) | 0.048 | 0.626<br>(0.385, 0.853) |

Analyses were adjusted for birth year, genotyping array, and first ten principal components. Odds ratios are given per standard deviation within ethnic group. Abbreviations: AAUC, covariate-adjusted area under the receiver-operator characteristics curve; CI, confidence interval; GWAS, genome-wide association study; PRS, polygenic risk score; SNP, single nucleotide polymorphism; AFR: African; EAS: East Asian; EUR: European, SAS: South Asian

The PRS-CS method usually outperformed the GWAS-hits-PRS in terms of association strength, accuracy and discrimination (Table 1). Especially for the breast cancer, the PRS-CS construct showed consistent effect sizes across the ancestry groups (1.66 ≤ OR [per unit S.D.] ≤ 1.77) and good discriminatory ability (0.64 ≤ AAUC ≤ 0.66)

To evaluate if the increased risk is observable with increasing score or only present in the tails of the distribution, we stratified the PRS, again standardized *within* each ancestry group, and detected a trend of increasing number of cases within the increasing PRS-CS score deciles. This trend was strikingly monotonous in the substantially larger sample of European ancestry and, except for the small sample of prostate cancer cases of East Asian ancestry, noticeable though more capricious in non-EA groups (Cochran-Armitage P < 0.00297; Fig 2, S3 and S4 Tables). We saw similar trends for the GWAS hit PRS (S1 Fig, S3 and S4 Tables).

**Fig 2.**
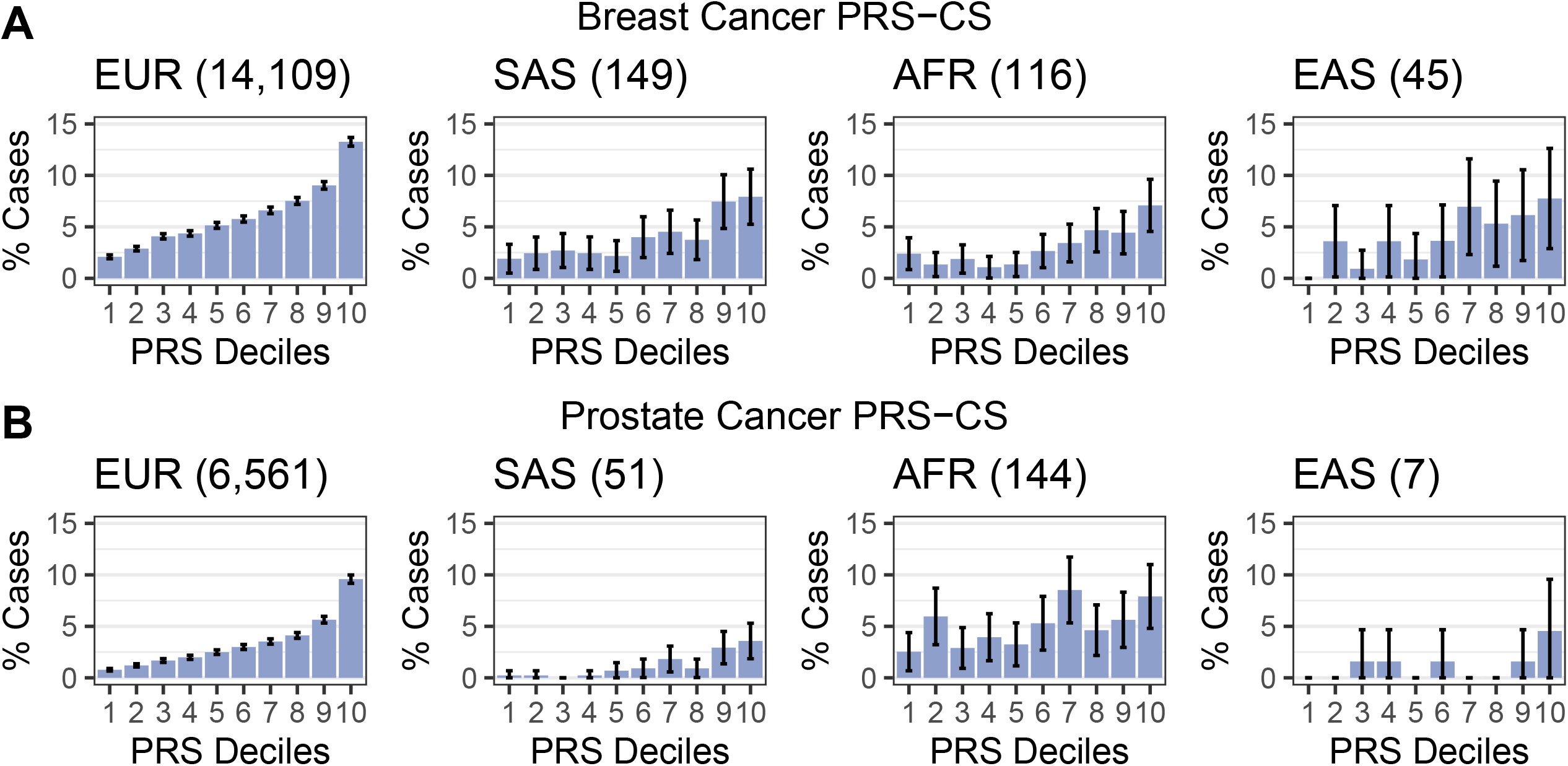
Observed case proportion across PRS-CS-based cancer PRS risk deciles. Proportions of breast cancer cases (A) and prostate cancer cases (B) stratified by ancestry groups are shown. Total case counts per ancestry group are given in parentheses. Underlying sample counts and corresponding Cochran-Armitage Test for Trend P-values are reported in S3 and S4 Tables.

Finally, we quantified the PRS’s ability to enrich cases in the top 10% of the PRS distribution (defined in controls *within* each ancestry group) when compared to the bottom 90%. We observed an enrichment for breast cancer cases in the tail of the PRS distribution when we defined the top 10% within each ancestry group (breast cancer: OR Top10% > 2.18; prostate cancer: OR Top10% > 1.41). The enrichment was particularly sizable for breast cancer PRS-CS score for cases in European and African ancestry females (OR Top10%: 2.81 [95% CI: 2.69, 2.93] and 2.88 [95% CI: 1.85, 4.48], respectively) as well as for the prostate cancer PRS-CS score for cases in European, South Asian and East Asian ancestry males (OR Top10%: 4.00 [95% CI: 3.78, 4.23]; 4.41 [95% CI: 2.43, 8.04] and 6.53 [95% CI: 1.71, 25.0], respectively; Table 2).

**Table 2.** Case enrichment in breast and prostate cancer PRS top 10% versus bottom 90%.

| GWAS Trait / Outcome | Ancestry Group | GWAS Hits PRS |  | PRS-CS PRS |  |
| --- | --- | --- | --- | --- | --- |
|  |  | OR Top 10% (95% CI) | P | OR Top 10% (95% CI) | P |
| Overall Breast Cancer | EUR | 2.36<br>(2.26, 2.47) | 1.0x10<sup>-328</sup> | 2.81<br>(2.69, 2.93) | 2.5x10<sup>-499</sup> |
|  | SAS | 2.54<br>(1.70, 3.79) | 5.98x10<sup>-6</sup> | 2.33<br>(1.55, 3.51) | 5.01x10<sup>-5</sup> |
|  | AFR | 2.18<br>(1.36, 3.49) | 0.00116 | 2.88<br>(1.85, 4.48) | 2.75x10<sup>-6</sup> |
|  | EAS | 3.52<br>(1.79, 6.9) | 0.000254 | 2.60<br>(1.25, 5.40) | 0.0106 |
| Prostate Cancer | EUR | 3.32<br>(3.13, 3.52) | 1.26x10<sup>-346</sup> | 4.00<br>(3.78, 4.23) | 3.77x10<sup>-495</sup> |
|  | SAS | 3.11<br>(1.66, 5.84) | 0.000409 | 4.41<br>(2.43, 8.04) | 1.18x10<sup>-6</sup> |
|  | AFR | 1.41<br>(0.85, 2.34) | 0.179 | 1.78<br>(1.09, 2.92) | 0.0223 |
|  | EAS | 4.89<br>(1.26, 19.0) | 0.0219 | 6.53<br>(1.71, 25.0) | 0.00614 |

Abbreviations: PRS, polygenic risk score; AFR, African; EAS, East Asian; EUR, European, SAS, South Asian

## Discussion

Overall, our findings in the UKB data are encouraging and suggest that cancer PRS derived from large EA-based GWAS can, to a certain degree, be useful for risk stratification *within* EA or *within* non-EA individuals even though their distributions are dissimilar. However, there are limitations in regard to the generalizability of this approach. First, a matching ancestry group with sufficiently large control sample sizes is needed to adequately place a person’s PRS within its reference PRS distributions. In this study, we obtained more homogenous groups by combining self-reported ethnic groups with genetically inferred ancestry groups. However, even within such groups an adjustment for any remaining population stratification, e.g., by including the first ten principal components, should be considered.

Secondly, overall breast and prostate cancer were selected because they offered several advantages compared to other traits: their estimated heritability is relatively high [17, 19, 20], they are common across all ancestry groups (breast cancer 3.1 – 6.2%; prostate cancer 1.2 – 5.1%; S1 Table) and each had summary statistics publicly available from large EA-based GWAS meta-analyses.

Thirdly, the UKB study individuals were recruited from the same country, the UK, where healthcare coverage and non-genetic risk factors might be more similar compared to diverse ancestries from geographically separate populations. Though we recognize that lifestyle, health disparities and socioeconomic factors (e.g., education and income, S1 Table) might vary between ethnic groups of the UKB study.

While a fraction of risk variants is likely population-specific, our observation of a decent predictive PRS performance across ancestry groups indicated that, for the two analyzed cancers, a fraction of the cancer risk variants obtained from an EA-based GWAS is shared with non-EA groups. So, while PRS that rely on EA-based GWAS were reported to be not ideal for non-EA groups, they can be useful for risk stratification also in non-EA groups. In our examples, the proportion of cases by PRS risk decile was informative within the studies ancestry group, i.e., an increasing PRS was associated with increased proportion of cases also among non-EA groups. However, we noted that the EA-based prostate cancer PRS performed particularly poor in AFR males indicating ancestry-specific diversity for prostate cancer as previously reported [21]. This also suggested that transferability of PRS across ancestries needs to be carefully evaluated by cancer and by ancestry group.

We recommend that PRS be constructed using GWAS based on the same ancestry group, if large diverse GWAS and their summary statistics are available. In the absence of large-scale GWAS for non-EA groups, several groups are developing methods to improve PRS performance in non-EA groups. These methods may leverage evidence that SNP selection based on EA-based GWAS is generally appropriate while the use of EA-based GWAS effect sizes in ethnically mismatched groups might not [22]. Duncan *et al*. [5] highlight the need for improved understanding and consideration of LD and variant frequencies when applying European ancestry based GWAS to non-EA groups, while at the same time calling for large-scale GWAS in diverse populations [5]. Modelling ancestry into polygenic risk predictors or focusing on global risk variants might allow the retention of comparable predictive power across ancestries [8] and allow risk stratification also in understudies populations as shown for Hispanics/Latinos [10]. However, a restriction to global risk variants, e.g., defined by similar frequencies across all ancestry groups, might lead to the exclusion of true causal risk variants. When we applied such a global risk variant approach to the current dataset through simple frequency filtering, we made PRS distributions more similar across ancestry groups but also observed markedly reduced predictive power (Figs S2-5). While efforts are underway to contribute more diverse samples to genetic studies, their sample sizes will trail behind sample sizes of European ancestry GWAS for a long time [6]. Multiethnic PRS that combine larger EA-based GWAS with smaller GWAS of the target ancestry group were recently proposed and might alleviate the discrepancies in sample sizes for the time being [23].

Taken together, our findings suggest that cross-ancestry cancer PRS can be useful for risk stratification, especially when there is a lack of well-powered diverse cancer GWAS. However, caution needs to be applied to the interpretation and application of such genetic risk predictors as they can be prone to multiple sources of bias [8].

## Materials and Methods

### Subjects / Genotypes

The UK Biobank (UKB) is a population-based cohort collected from multiple sites across the United Kingdom and includes over 500,000 participants aged between 40 and 69 years when recruited in 2006–2010 [15]. The open-access UK Biobank data used in this study included questionnaire data, electronic health record data, and genotype and genotyped derived data. UK Biobank received ethical approval from the NHS National Research Ethics Service North West (11/NW/0382). The present analyses were conducted under UK Biobank data application number 24460.

We excluded 2,338 samples which were flagged by the UK Biobank quality control documentation as (1) het.missing.outliers, (2) putative.sex.chromosome.aneuploidy, (3) excess.relatives, (4) excluded.from.kinship.inference, (5) the reported gender did not match the inferred sex, (6) withdrew from the UKB study and (7) were not included in the phased and imputed genotype data of chromosomes 1-22, and X (in.Phasing.Input.chr1_22 and in.Phasing.Input.chrX) [24]. 485,434 individuals remained after sample QC filtering. We used the UK BioBank Imputed Dataset (v3, https://www.ebi.ac.uk/ega/datasets/EGAD00010001474) and limited analyses to variants with imputation information score >= 0.3 and MAF >= 0.01%.

### Phenotype and covariate data

For the current study we included self-reported ethnic group (field: 21000), sex (fields: 31, 22001), income (field: 738), education (field: 6138), diet (fields: 1309, 1319, 1329, 1339, 1349, 1359, 1369, 1379, 1389), year of birth (field: 34).

We used ICD9 (fields: 40013, 41203, and 41205) and ICD10 code data (fields: 40001, 40002, 40006, 41201, 41202, and 41204) to define breast and prostate cancer case control studies using PheWAS codes ‘174.1’ and ‘185’ [25]. Underlying ICD codes for cases were as follows: breast cancer: ICD9: 233.0; ICD10: C50.*, D05.1, D05.7, D05.9, and Z85.3; and prostate cancer: ICD9: 185, 233.4; ICD10: C61, D07.5.

We used both principal component-based ancestry prediction and self-reported ethnic information to define ancestry groups. For the ancestry prediction, we applied online augmentation, decomposition and Procrustes (OADP) method to the genotype data of 488,366 UK Biobank samples with 2492 samples from the 1000 Genomes Project data as the reference (FRAPOSA; see **Web Resources**)[26] to infer the super populations membership (AFR: African, AMR: Ad Mixed American, EAS: East Asian, EUR: European, and SAS: South Asian ancestry). We combined the self-reported ethnic group and the inferred super population membership to define the following four ancestry groups for downstream analyses: African (self-reported “Black or Black British” and inferred AFR), East Asian (self-reported “Asian or Asian British” or East Asian and inferred EAS), European (self-reported European and inferred EUR), and South Asian individuals (self-reported “Asian or Asian British” and inferred SAS). By doing so we excluded individuals with admixed and/or unknown ancestry as well as individuals where self-reported ethnic group did not match their inferred ancestry.

For each cancer trait and each ancestry group, we extracted a maximal set of unrelated individuals (defined as kinship coefficient < 0.0884) [27] by first selecting a maximal set of unrelated cases before selecting a set of unrelated controls that was not related to any of the selected cases. [28]

### PRS Construction

PRS combine information across a defined set of genetic loci, incorporating each locus’s association with the target trait. The PRS for person j takes the form PRS_*j*_=∑_*i*_ *β*_*i*_*G*_*ij*_ where *i* indexes the included loci for that trait, weight *β*_*i*_ is the log odds ratios retrieved from the external GWAS summary statistics for locus *i*, and *G*_*ij*_ is a continuous version of the measured dosage data for the risk allele on locus *i* in subject *j*.

We downloaded full GWAS summary statistics made available by the “Breast Cancer Association Consortium” (BCAC) [20], and the “Prostate Cancer Association Group to Investigate Cancer Associated Alterations in the Genome” (PRACTICAL) [17] (also see **Web Resources**) both based on European ancestry samples. For each set of GWAS summary statistics, we create two PRS. For the first PRS construction method, we performed linkage disequilibrium (LD) clumping of variants with p-values below 5×10^−8^ by using the imputed allele dosages of 10,000 randomly selected samples and a pairwise correlation cut-off at r^2^ < 0.1 within 1Mb window. Using the resulting loci (“independent GWAS hits”), we calculated the weighted PRS (see above) denoted as “GWAS hits PRS”. For the second PRS construction method, we used the software package “PRS-CS” [18] to define a PRS based on the continuous shrinkage (CS) priors. PRS-CS uses a precomputed LD reference panel based on external European samples of the 1000 Genomes Project (“EUR reference”). We applied a MAF filter of 1 % and, in contrast to the GWAS Hits PRS only included autosomal variants that overlap between summary statistics, LD reference panel, and target panel. Full list of weights can be downloaded from our web site (see **Web Resources**).

We obtained deep sequenced data on the 2504 samples in the 1000 Genomes Project’s phase three panel that were generated by the New York Genome Center (see **Web Resources**). Sequencing data was filtered to have a minimum depth of 10, to be polymorphic and located on chromosomes 1 – 22, X. We stratified the data according to their super populations (AFR, African; AMR, Ad Mixed American; EAS, East Asian; EUR, European; SAS, South Asian) and calculated their population specific allele frequencies using PLINK 1.9 (see Web Resources). We created five sets of variants whose MAF was >1 % in AFR, EAS, EUR, SAS and whose maximal allele frequency difference between any of the four populations was below 5, 10, 15, 20 or 25%. The resulting sets were used to filter the GWAS summary statistics before running PRS-CS.

Using the R package “Rprs” (see **Web Resources**) and the weights from the two PRS methods, the dosage-based value of each PRS was then calculated for each UKB individual. For comparability of association effect sizes corresponding to the continuous PRS across cancer traits and PRS construction methods, we centered PRS values to their mean and scaled them to have a standard deviation of 1.

### Statistical Tests

For the PRS evaluations, we fit the following model for each PRS and cancer phenotype adjusting for covariates Birthyear, genotyping Array, and the first ten principal components (PC) using a complete case analysis:

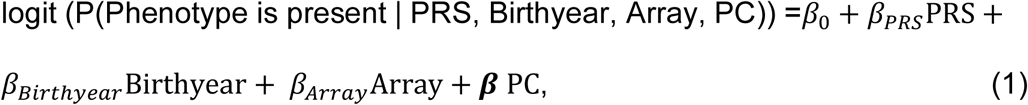

where the PCs were the first ten principal components obtained from the principal component analysis provided by the UK Biobank study and where “Array” represents the genotyping array.

For each PRS derived for each GWAS source/method combination, we also assessed the following PRS performance measures relative to observed binary disease status: overall association and the ability to discriminate between cases and controls as measured by the area under the covariate-adjusted receiver operating characteristic (AROC; semiparametric frequentist inference [29]) curve (denoted AAUC) using R package “ROCnReg” [30]. Firth’s bias reduction method was used to resolve the problem of separation in logistic regression (R package “brglm2”)[31, 32].

For each ancestry group (African, East Asian, European, and South Asian), we also stratified the UKB control dataset (i.e., the corresponding gender subset depending on cancer type) into ten groups of equal size by PRS deciles and determined the number of observed case subjects that were observed in the range of each risk decile. To assess for the presence of an association between cancer and increasing PRS risk deciles, we performed a Cochran Armitage Test for Trend implemented in the R package “DescTools” [33]. To study the ability of the PRS to identify high risk patients, we fit the above model (equation 1) by replacing the PRS with an indicator for whether the PRS value was in the top decile or not.

To test if the PRS means between the ancestry groups are equal we used ANOVA adjusting for genotyping array, birthyear and the first 10 principal components.

We used the STREGA checklist when writing our report [34]

## Supporting information

Completed STREGA Checklist

## Data Availability

Data cannot be shared publicly due to patient confidentiality. The data underlying the results presented in the study are available from the UK Biobank for researchers who meet the criteria for
access to confidential data.

http://www.ukbiobank.ac.uk/register-apply/

## Acknowledgement

This research has been conducted using the UK Biobank Resource under application number 24460. Additional acknowledgements of GWAS sources are listed in S1 Text.

## Funding

This material is based in part upon work supported by the National Institutes of Health/NIH (NCI P30CA046592 [LGF, MS, BM]), by the University of Michigan (UM-Precision Health Investigators Award U063790 [LGF, SP, YM, BM]), by the National Research Foundation of Korea (BP+ Program [SL]) and by the National Science Foundation under grant number DMS-1712933. Any opinions, findings, and conclusions or recommendations expressed in this material are those of the author(s) and do not necessarily reflect the views of the National Science Foundation.

## Web Resources

UK Biobank dataset, https://www.ebi.ac.uk/ega/datasets/EGAD00010001474

PubMed, https://www.ncbi.nlm.nih.gov/pubmed

FRAPOSA, https://github.com/daviddaiweizhang/fraposa

The Prostate Cancer Association Group to Investigate Cancer Associated Alterations in the Genome (PRACTICAL), http://practical.icr.ac.uk/blog/?page_id=8164

The Breast Cancer Association Consortium, http://bcac.ccge.medschl.cam.ac.uk/bcacdata/oncoarray/oncoarray-and-combined-summary-result/gwas-summary-results-breast-cancer-risk-2017/

PRS-CS, https://github.com/getian107/PRScs

Weights for constructed PRS, https://www.dropbox.com/sh/mwo23qhhlq42odw/AACCRQBsaNORBmnngN1U-wkwa

Deep sequenced 1000 Genomes Project data, ftp://ftp.1000genomes.ebi.ac.uk/vol1/ftp/data_collections/1000G_2504_high_coverage/working/

PLINK 1.9, https://www.cog-genomics.org/plink/

R package “Rprs”, https://github.com/statgen/Rprs

## Supporting information

**S1 Fig. Observed case proportion across GWAS hits-based cancer PRS risk deciles**.

**S2 Fig. Breast cancer PRS (PRS-CS) distributions before and after defining global risk variants**.

**S3 Fig. Prostate cancer PRS (PRS-CS) distributions before and after defining global risk variants**.

**S4 Fig. Breast cancer PRS (PRS-CS) associations based on unfiltered and five global risk variant sets**.

**S5 Fig. Prostate cancer PRS (PRS-CS) associations based on unfiltered and five global risk variant sets**.

**S1 Table. Demographics of the UK Biobank study**.

**S2 Table. Comparison of breast cancer and prostate cancer PRS stratified by ancestry group**. ANOVA test was adjusted using birth year, genotyping array and first ten principal components.

**S3 Table. Breast cancer PRS risk deciles calculated within females of each ancestry group**. Counts by ancestry group and case-control status.

**S4 Table. Prostate cancer PRS risk deciles calculated within males of each ancestry group**. Counts by ancestry group and case-control status.

**S1 Text. Supplemental Acknowledgements**.

## Notes

### Competing Interest Statement

The authors have declared no competing interest.

### Author Declarations

The open-access UK Biobank data used in this study included questionnaire data, electronic health record data, and genotype and genotyped derived data. UK Biobank received ethical approval from the NHS National Research Ethics Service North West (11/NW/0382).

