## Supplemental Information for "On Cross-ancestry Cancer Polygenic Risk Scores"

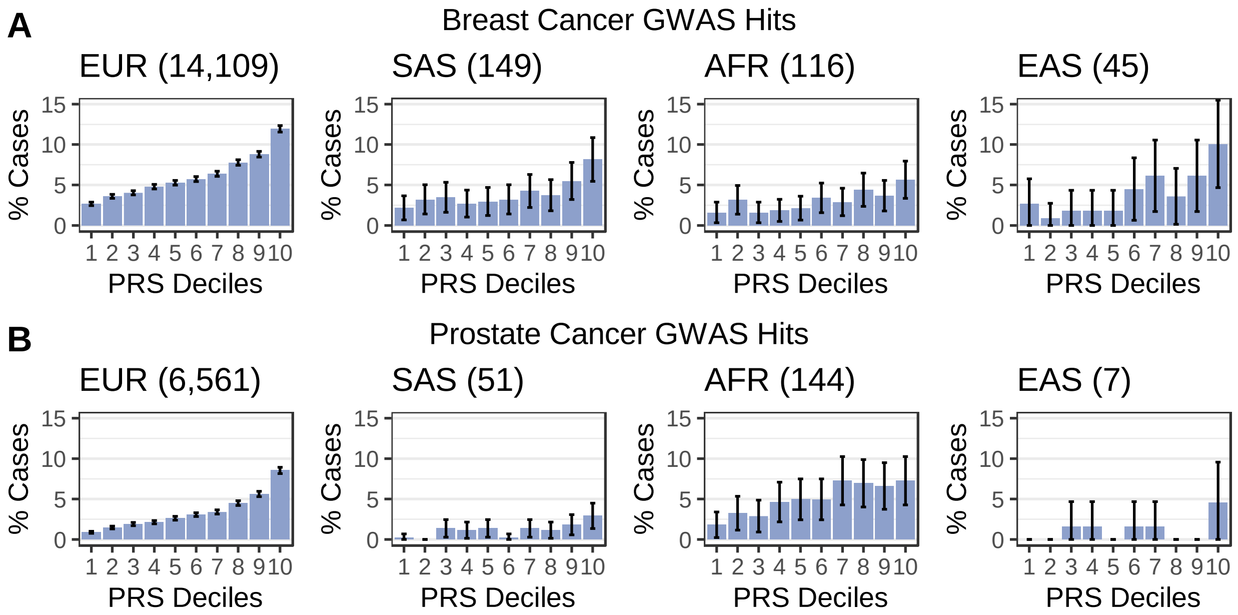


**S1 Figure. Observed case proportion across GWAS hits-based cancer PRS risk deciles**. Proportions of breast cancer cases (A) and prostate cancer cases (B) stratified by ancestry groups are shown. Total case counts per ancestry group are given in parentheses. Underlying sample counts and corresponding Cochran-Armitage Test for Trend P-values are reported in S3 and S4 Tables. Abbreviations: AFR: African; EAS: East Asian; EUR: European, SAS: South Asian.

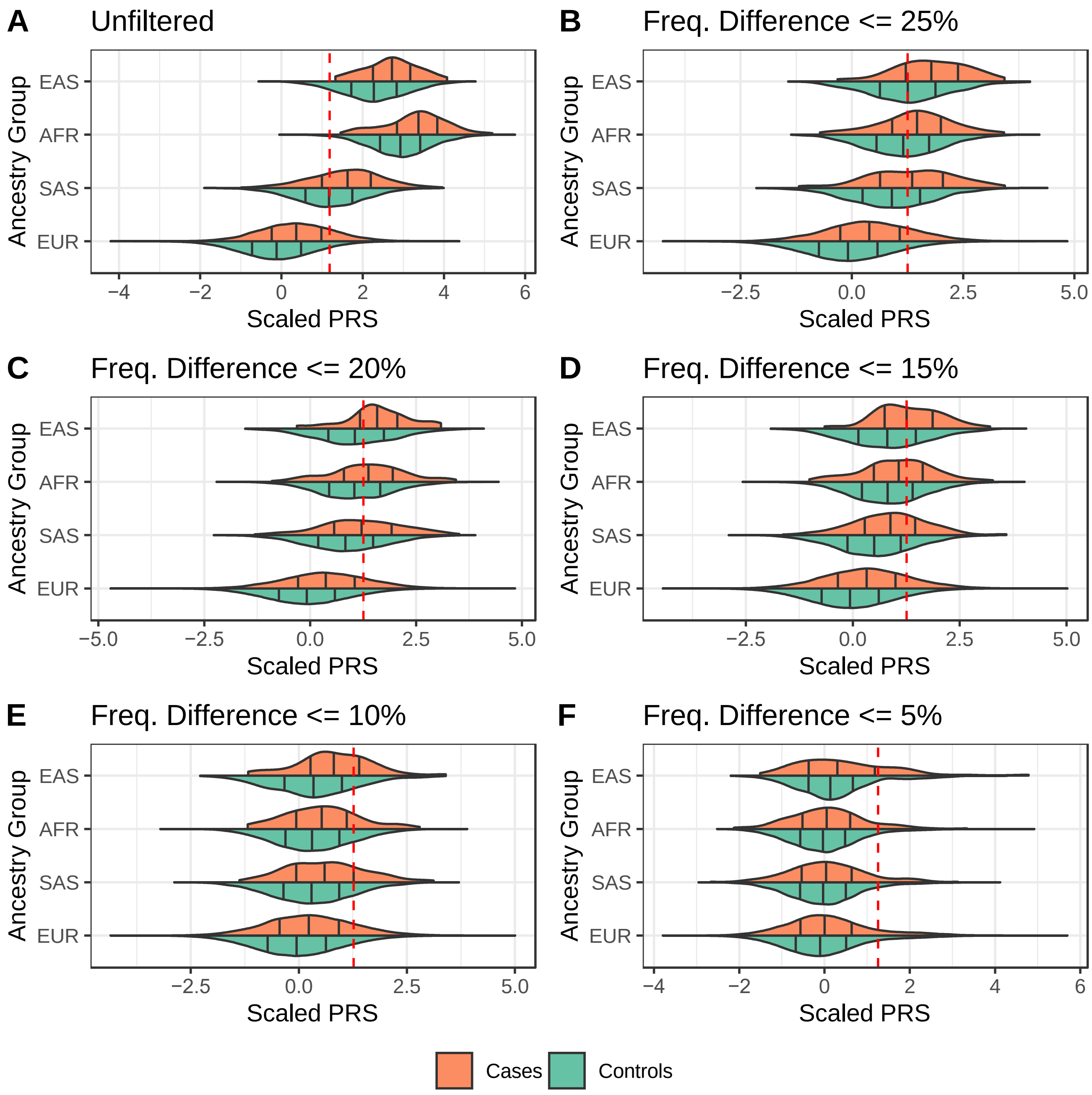


**S2 Figure. Breast cancer PRS (PRS-CS) distributions before (A) and after (B – F) defining global risk variants.** Five sets of global variants were defined as variant whose allele frequency differences between the four ancestry groups within the 1000 Genomes Project reference were below 25% (B), 20% (C), 15% (D), 10 % (E) and 5 % (F). Red lines indicate 10% quantiles of the corresponding UKB PRS distribution in all controls. Abbreviations: AFR: African; EAS: East Asian; EUR: European, SAS: South Asian

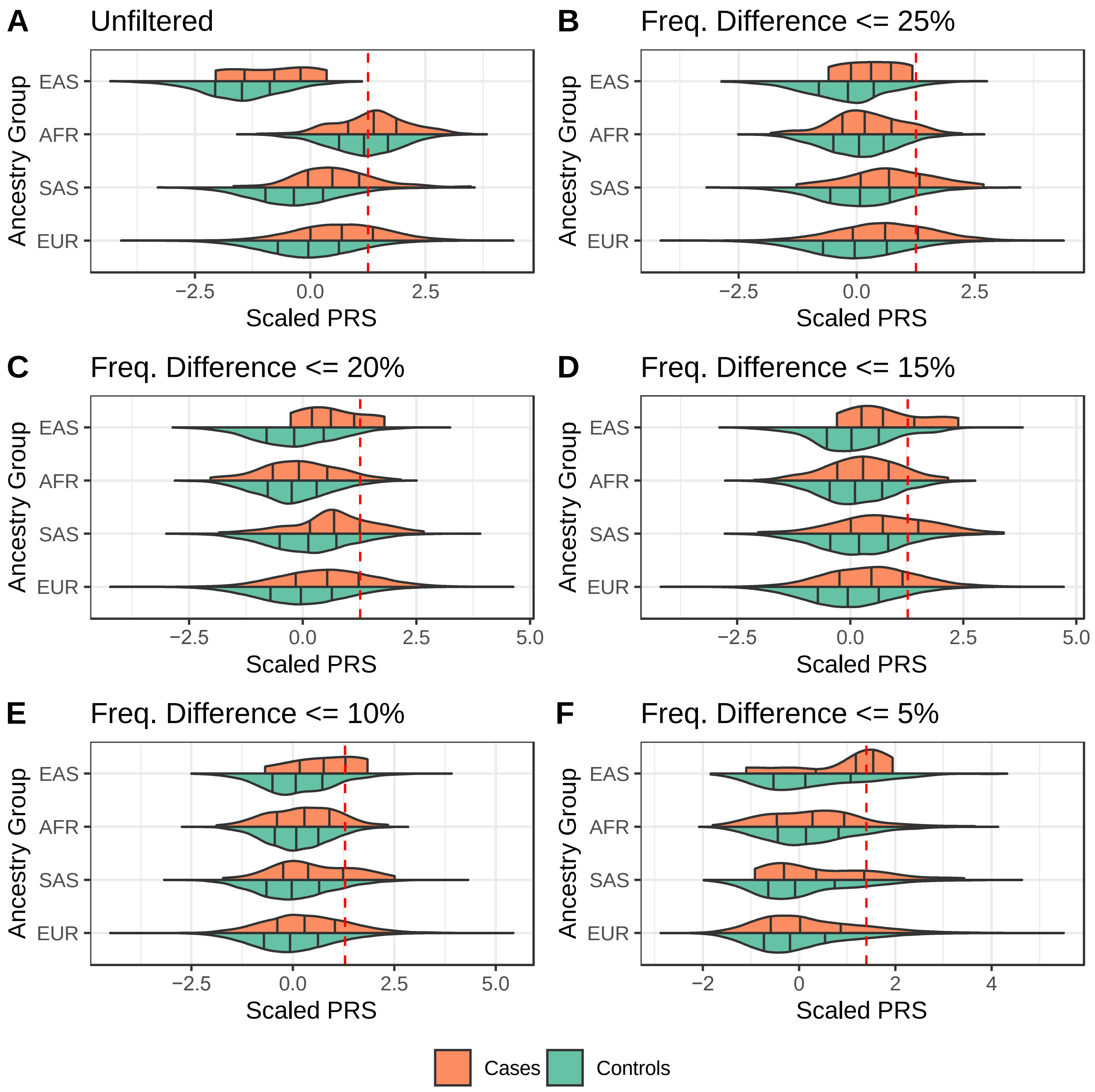


**S3 Figure. Prostate cancer PRS (PRS-CS) distributions before (A) and after (B – F) defining global risk variants.** Five sets of global variants were defined as variant whose allele frequency differences between the four ancestry groups within the 1000 Genomes Project reference were below 25% (B), 20% (C), 15% (D), 10 % (E) and 5 % (F). Red lines indicate 10% quantiles of the corresponding UKB PRS distribution in all controls. Abbreviations: AFR: African; EAS: East Asian; EUR: European, SAS: South Asian

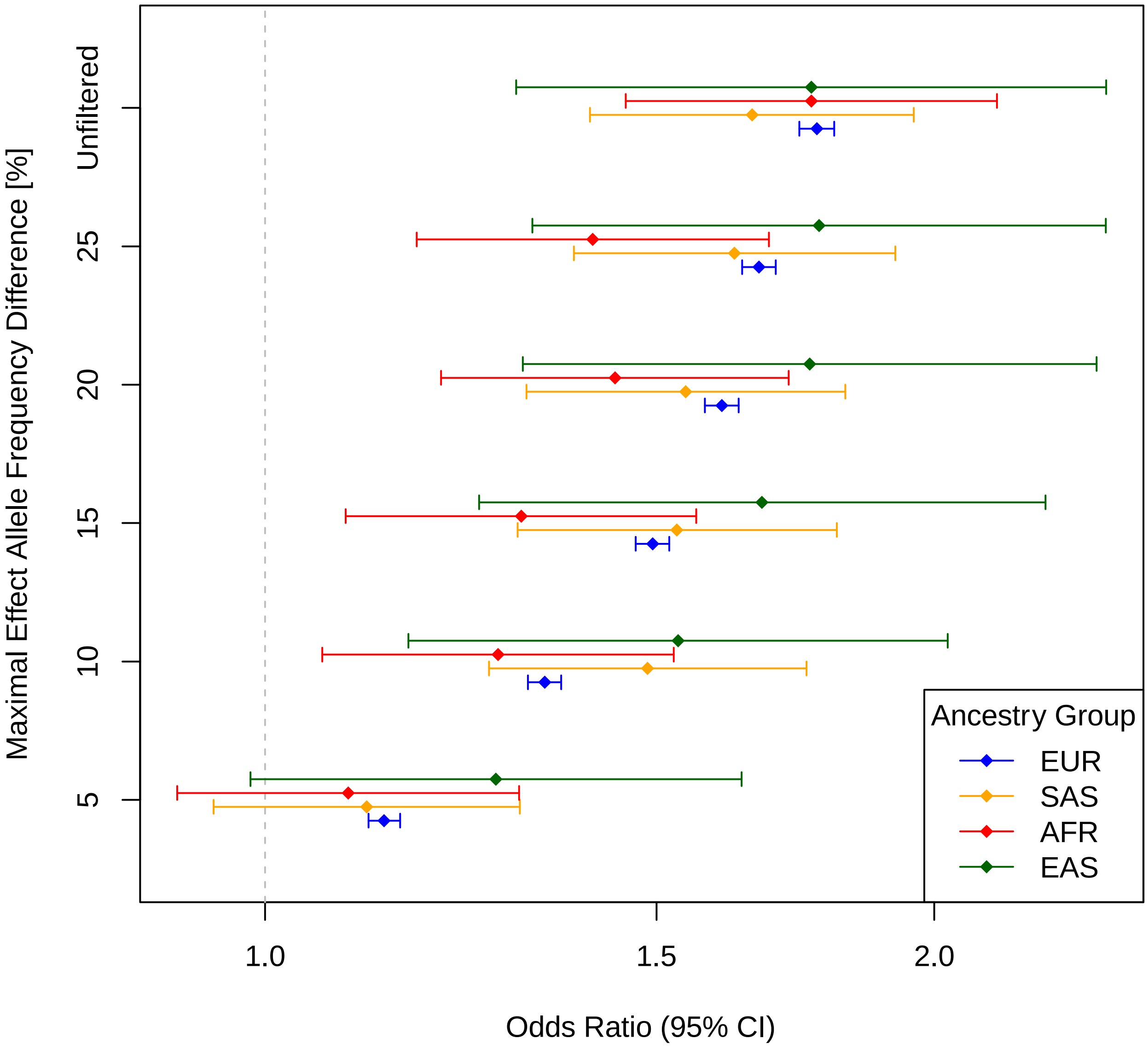


**S4 Figure. Breast cancer PRS (PRS-CS) associations based on unfiltered and five global risk variant sets.** Associations are shown across the ancestry groups. Analyses were adjusted for birth year, genotyping array, and first ten principal components. Five sets of global variants were defined as variant whose allele frequency differences between the four ancestry groups within the 1000 Genomes Project reference were below 5, 10, 15, 20 and 25%. Abbreviations: AFR: African; EAS: East Asian; EUR: European, SAS: South Asian

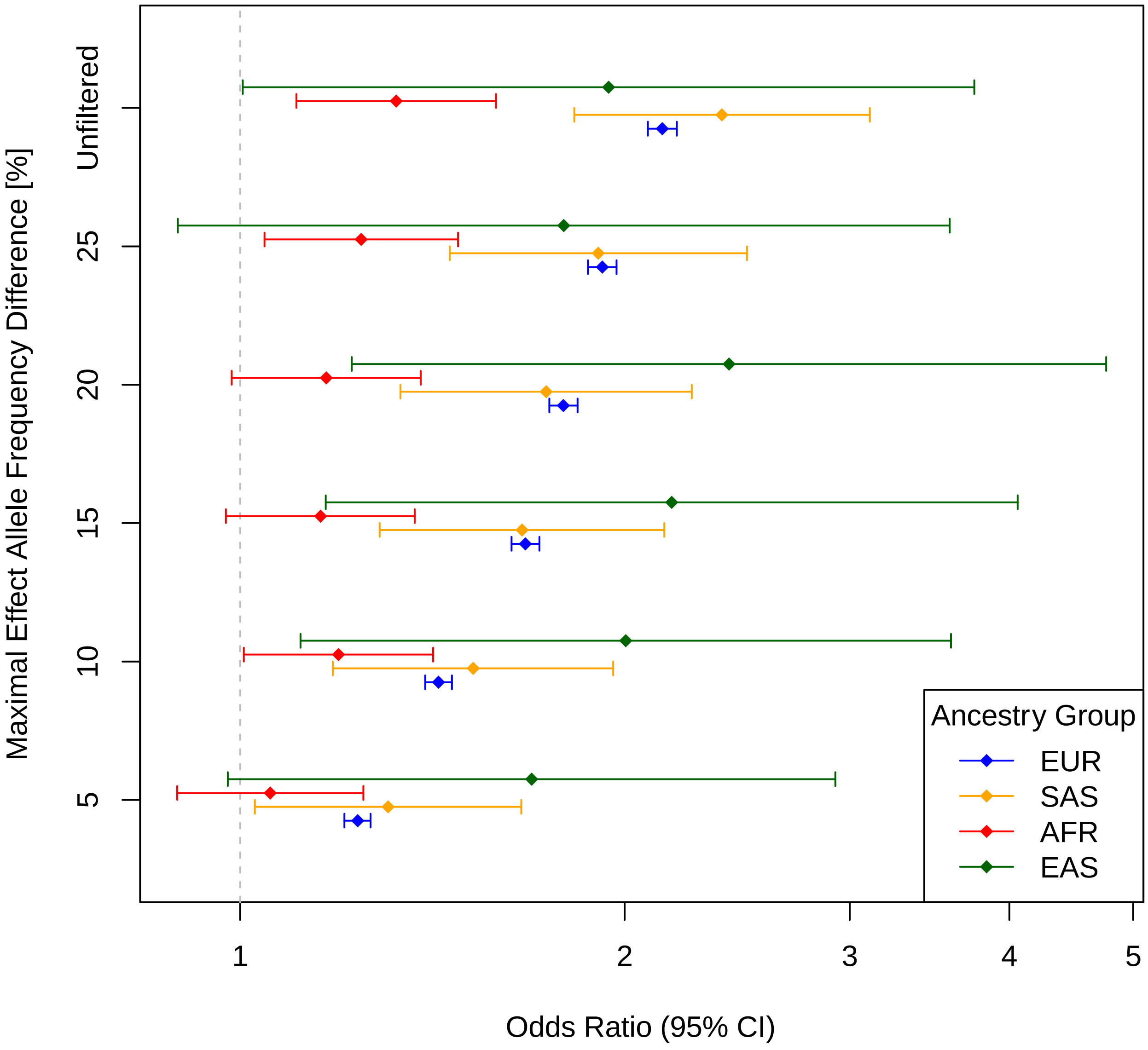


**S5 Figure. Prostate cancer PRS (PRS-CS) associations based on unfiltered and five global risk variant sets.** Associations are shown across the ancestry groups. Analyses were adjusted for birth year, genotyping array, and first ten principal components. Five sets of global variants were defined as variant whose allele frequency differences between the four ancestry groups within the 1000 Genomes Project reference were below 5, 10, 15, 20 and 25%. Abbreviations: AFR: African; EAS: East Asian; EUR: European, SAS: South Asian

**S1 Table.** Demographics of the UK Biobank study

| | Ancestry Group[c] | | | |
|---|---|---|---|---|
| | **EUR** | **SAS** | **AFR** | **EAS** |
| **n** | 417423 | 8103 | 6607 | 1743 |
| **Males (%)** | 189151 (45.3) | 4356 (53.8) | 2825 (42.8) | 629 (36.1) |
| **Breast Cancer (%)[a]** | 14109 (6.2) | 149 (4.0) | 116 (3.1) | 45 (4.0) |
| **Prostate Cancer (%)[b]** | 6561 (3.5) | 51 (1.2) | 144 (5.1) | 7 (1.1) |
| **Education (%)** | | | | |
| College or University degree | 134688 (32.3) | 3137 (38.9) | 2196 (33.5) | 827 (47.5) |
| A levels/AS levels or equivalent | 46984 (11.3) | 704 (8.7) | 449 (6.8) | 129 (7.4) |
| O levels/GCSEs or equivalent | 89328 (21.5) | 1291 (16.0) | 1137 (17.3) | 202 (11.6) |
| CSEs or equivalent | 22644 (5.4) | 451 (5.6) | 506 (7.7) | 44 (2.5) |
| NVQ or HND or HNC or equivalent | 27722 (6.7) | 378 (4.7) | 725 (11.1) | 78 (4.5) |
| Other professional qualifications e.g., nursing, teaching | 22345 (5.4) | 372 (4.6) | 509 (7.8) | 129 (7.4) |
| None of the above | 69081 (16.6) | 1286 (16.0) | 825 (12.6) | 233 (13.4) |
| Prefer not to answer | 3628 (0.9) | 435 (5.4) | 214 (3.3) | 98 (5.6) |
| **Income (%)** | | | | |
| Greater than 100,000 | 21016 (5.0) | 378 (4.7) | 75 (1.1) | 86 (4.9) |
| 52,000 to 100,000 | 76770 (18.4) | 1066 (13.2) | 613 (9.3) | 238 (13.7) |
| 31,000 to 51,999 | 95850 (23.0) | 1255 (15.6) | 1174 (17.9) | 343 (19.7) |
| 18,000 to 30,999 | 90801 (21.8) | 1426 (17.7) | 1501 (22.9) | 307 (17.6) |
| Less than 18,000 | 76923 (18.5) | 1915 (23.8) | 1719 (26.2) | 397 (22.8) |
| Do not know | 16180 (3.9) | 695 (8.6) | 685 (10.4) | 121 (7.0) |
| Prefer not to answer | 38880 (9.3) | 1319 (16.4) | 794 (12.1) | 248 (14.3) |

[a] females only; [b] males only; [c] AFR: African; EAS: East Asian; EUR: European, SAS: South Asian

**S2 Table** Comparison of breast cancer and prostate cancer PRS stratified by ancestry group. ANOVA test was adjusted using birth year, genotyping array and first ten principal components.

| | Ancestry Group[c] | n | GWAS Hits PRS | | PRS-CS PRS | |
|---|---|---|---|---|---|---|
| | | | Mean (s.d.) | ANOVA P (F-value) | Mean (s.d.) | ANOVA P (F-value) |
| Breast Cancer PRS [a] | EUR | 228,272 | -0.0366 (0.985) | $4.6 \times 10^{-2148}$ (3369.1) | -0.0794 (0.903) | $1.67 \times 10^{-11149}$ (19114.3) |
| | SAS | 3,747 | 0.539 (0.898) | | 1.19 (0.853) | |
| | AFR | 3,782 | 1.37 (0.713) | | 2.94 (0.74) | |
| | EAS | 1,114 | 1.05 (0.776) | | 2.29 (0.795) | |
| Prostate Cancer PRS [b] | EUR | 189,151 | -0.00601 (1.01) | $2.31 \times 10^{-141}$ (218.26) | -0.00509 (0.99) | $3.73 \times 10^{-1229}$ (1915.9) |
| | SAS | 4,356 | -0.0145 (0.863) | | -0.32 (0.923) | |
| | AFR | 2,825 | 0.468 (0.666) | | 1.16 (0.771) | |
| | EAS | 629 | -0.194 (0.757) | | -1.45 (0.882) | |

Abbreviations: GWAS, genome-wide association study; PRS, polygenic risk score; PRS-CS, PRS method based on the continuous shrinkage (CS) priors; PRS, polygenic risk score; s.d., standard deviation
[a] female individuals only; [b] male individuals only; [c] AFR: African; EAS: East Asian; EUR: European, SAS: South Asian

**S3 Table.** Breast cancer PRS risk deciles calculated within females of each ancestry group. Counts by ancestry group and case-control status.

| Ancestry Group[a] | | Risk Decile | | | | | | | | | | |
|---|---|---|---|---|---|---|---|---|---|---|---|---|
| | | **1** | **2** | **3** | **4** | **5** | **6** | **7** | **8** | **9** | **10** | **Cochran-Armitage Test for Trend P** |
| **GWAS Hits PRS** | | | | | | | | | | | | |
| **EUR** | Cases | 588 | 800 | 903 | 1079 | 1194 | 1305 | 1460 | 1805 | 2069 | 2906 | 6.38e-551 |
| | Controls | 21417 | 21416 | 21416 | 21416 | 21417 | 21416 | 21416 | 21416 | 21416 | 21417 | |
| **SAS** | Cases | 8 | 12 | 13 | 10 | 11 | 12 | 16 | 14 | 21 | 32 | 2.45e-05 |
| | Controls | 360 | 360 | 360 | 359 | 360 | 360 | 359 | 360 | 360 | 360 | |
| **AFR** | Cases | 6 | 12 | 6 | 7 | 8 | 13 | 11 | 17 | 14 | 22 | 0.000258 |
| | Controls | 367 | 367 | 366 | 367 | 366 | 367 | 366 | 367 | 366 | 367 | |
| **EAS** | Cases | 3 | 1 | 2 | 2 | 2 | 5 | 7 | 4 | 7 | 12 | 0.000133 |
| | Controls | 107 | 107 | 107 | 107 | 107 | 106 | 107 | 107 | 107 | 107 | |
| **PRS-CS PRS** | | | | | | | | | | | | |
| **EUR** | Cases | 457 | 636 | 911 | 977 | 1162 | 1310 | 1514 | 1741 | 2126 | 3275 | 6.43e-758 |
| | Controls | 21417 | 21416 | 21416 | 21416 | 21417 | 21416 | 21416 | 21416 | 21416 | 21417 | |
| **SAS** | Cases | 7 | 9 | 10 | 9 | 8 | 15 | 17 | 14 | 29 | 31 | 1.45e-08 |
| | Controls | 360 | 360 | 360 | 359 | 360 | 360 | 359 | 360 | 360 | 360 | |
| **AFR** | Cases | 9 | 5 | 7 | 4 | 5 | 10 | 13 | 18 | 17 | 28 | 5.92e-08 |
| | Controls | 367 | 367 | 366 | 367 | 366 | 367 | 366 | 367 | 366 | 367 | |
| **EAS** | Cases | 0 | 4 | 1 | 4 | 2 | 4 | 8 | 6 | 7 | 9 | 0.000342 |
| | Controls | 107 | 107 | 107 | 107 | 107 | 106 | 107 | 107 | 107 | 107 | |

[a] AFR: African; EAS: East Asian; EUR: European, SAS: South Asian

**S4 Table.** Prostate cancer PRS risk deciles calculated within males of each ancestry group. Counts by ancestry group and case-control status.

| Ancestry Group[a] | | Risk Decile | | | | | | | | | | |
|---|---|---|---|---|---|---|---|---|---|---|---|---|
| | | 1 | 2 | 3 | 4 | 5 | 6 | 7 | 8 | 9 | 10 | Cochran-Armitage Test for Trend P |
| **GWAS Hits PRS** | | | | | | | | | | | | |
| **EUR** | Cases | 164 | 275 | 355 | 399 | 492 | 579 | 644 | 860 | 1088 | 1705 | 2.62e-507 |
| | Controls | 18259 | 18259 | 18259 | 18259 | 18259 | 18259 | 18259 | 18259 | 18259 | 18259 | |
| **SAS** | Cases | 1 | 0 | 6 | 5 | 6 | 1 | 6 | 5 | 8 | 13 | 0.000144 |
| | Controls | 431 | 430 | 431 | 430 | 431 | 430 | 430 | 431 | 430 | 431 | |
| **AFR** | Cases | 5 | 9 | 8 | 13 | 14 | 14 | 21 | 20 | 19 | 21 | 2.41e-05 |
| | Controls | 269 | 268 | 268 | 268 | 268 | 268 | 268 | 268 | 268 | 268 | |
| **EAS** | Cases | 0 | 0 | 1 | 1 | 0 | 1 | 1 | 0 | 0 | 3 | 0.134 |
| | Controls | 63 | 62 | 62 | 62 | 62 | 62 | 62 | 62 | 62 | 63 | |
| **PRS-CS PRS** | | | | | | | | | | | | |
| **EUR** | Cases | 144 | 223 | 312 | 371 | 467 | 564 | 668 | 784 | 1092 | 1936 | 6.6e-630 |
| | Controls | 18259 | 18259 | 18259 | 18259 | 18259 | 18259 | 18259 | 18259 | 18259 | 18259 | |
| **SAS** | Cases | 1 | 1 | 0 | 1 | 3 | 4 | 8 | 4 | 13 | 16 | 2.76e-10 |
| | Controls | 431 | 430 | 431 | 430 | 431 | 430 | 430 | 431 | 430 | 431 | |
| **AFR** | Cases | 7 | 17 | 8 | 11 | 9 | 15 | 25 | 13 | 16 | 23 | 0.00297 |
| | Controls | 269 | 268 | 268 | 268 | 268 | 268 | 268 | 268 | 268 | 268 | |
| **EAS** | Cases | 0 | 0 | 1 | 1 | 0 | 1 | 0 | 0 | 1 | 3 | 0.0787 |
| | Controls | 63 | 62 | 62 | 62 | 62 | 62 | 62 | 62 | 62 | 63 | |

[a] AFR: African; EAS: East Asian; EUR: European, SAS: South Asian

### S1 Text. Supplemental Acknowledgements

Breast Cancer Association Consortium (BCAC)

The breast cancer genome-wide association analyses were supported by the Government of Canada through Genome Canada and the Canadian Institutes of Health Research, the 'Ministère de l'Économie, de la Science et de l'Innovation du Québec' through Genome Québec and grant PSR-SIIRI-701, The National Institutes of Health (U19 CA148065, X01HG007492), Cancer Research UK (C1287/A10118, C1287/A16563, C1287/A10710) and The European Union (HEALTH-F2-2009-223175 and H2020 633784 and 634935). All studies and funders are listed in Michailidou et al (2017).

Prostate Cancer Association Group to Investigate Cancer Associated Alterations in the Genome (PRACTICAL)

The Prostate cancer genome-wide association analyses are supported by the Canadian Institutes of Health Research, European Commission's Seventh Framework Programme grant agreement n° 223175 (HEALTH-F2-2009-223175), Cancer Research UK Grants C5047/A7357, C1287/A10118, C1287/A16563, C5047/A3354, C5047/A10692, C16913/A6135, and The National Institute of Health (NIH) Cancer Post-Cancer GWAS initiative grant: No. 1 U19 CA 148537-01 (the GAME-ON initiative).

Genotyping of the OncoArray was funded by the US National Institutes of Health (NIH) [U19 CA 148537 for ELucidating Loci Involved in Prostate cancer SuscEptibility (ELLIPSE) project and X01HG007492 to the Center for Inherited Disease Research (CIDR) under contract number HHSN268201200008I] and by Cancer Research UK grant

A8197/A16565. Additional analytic support was provided by NIH NCI U01 CA188392 (PI: Schumacher).

We would also like to thank the following for funding support: The Institute of Cancer Research and The Everyman Campaign, The Prostate Cancer Research Foundation, Prostate Research Campaign UK (now PCUK), The Orchid Cancer Appeal, Rosetrees Trust, The National Cancer Research Network UK, The National Cancer Research Institute (NCRI) UK. We are grateful for support of NIHR funding to the NIHR Biomedical Research Centre at The Institute of Cancer Research and The Royal Marsden NHS Foundation Trust.
